# An Experimental Investigation of the Relationship between AI-Human Workflow Design and Legal Liability for Radiologists: The Erroneous-Change Penalty and Omission Bias

**DOI:** 10.64898/2026.05.20.26353717

**Authors:** Elizabeth C. Song, Michael H. Bernstein, Brian Sheppard, Michael A. Bruno, Grayson L. Baird

## Abstract

**Background:** With growing impetus to integrate artificial intelligence (AI) tools into radiology, clinical practices must navigate workflow redesign. This carries implications for medical malpractice liability.

**Methods:** We conducted an online vignette experiment with United States adults who acted as hypothetical jurors in a malpractice case involving a missed intracranial hemorrhage. Participants (n=2,347) were randomized to one of 22 conditions: a no-AI control and 21 conditions involving a hypothetical AI system. These twenty-one conditions varied by whether (1) a single-read or double-read workflow was used, (2) the radiologist’s initial interpretation was documented, (3) the radiologist changed their interpretation after viewing AI output, (4) the AI detected the abnormality, and (5) the AI error rate—False Discovery Rate (FDR) or False Omission Rate (FOR)—was provided to participants only, both participants and radiologist, or neither. The primary outcome was perceived liability, assessed by whether the radiologist met their duty of care.

**Findings:** Perceived liability differed across conditions (p<0.0001). Double-read workflows (p<0.0001), documenting initial interpretations (p=0.0125), and providing participants with AI error rates, including the FDR (p=0.0038) or FOR (p=0.0035), reduced perceived liability. Liability was also lower when AI was incorrect (p<0.0001). Radiologists’ awareness of AI error rates did not significantly impact liability. Notably, we observed an “erroneous change penalty”: the greatest liability occurred when radiologists initially identified an abnormality but later changed their interpretation to normal after seeing that AI identified the case as normal; conversely, perceived liability was lowest with documented, double-read workflows.

**Interpretation:** Double-read workflows with documented initial interpretations and disclosure of AI error rates reduce perceived liability, though changing a correct initial interpretation increases it. Strategic workflow design is critical for successful AI implementation that can mitigate malpractice risk.

**RESEARCH IN CONTEXT:** *Evidence before this study:* Emerging research has identified several factors that shape how artificial intelligence (AI) systems are integrated into clinical workflows. Beyond technical performance, factors such as disease prevalence, documentation practices, and workflow design have all been shown to play a role in the implementation of AI tools and how they will inevitably affect physician liability. Vignette experiments have separately identified cognitive biases and mitigating measures that can be integrated into workflow design, such as disclosing AI error rates and documenting independent interpretations before reviewing AI output.

*Added value of this study:* This study builds on prior work by examining how multiple aspects of AI implementation influence perceptions of legal liability when hypothetical jurors are asked to adjudicate a medical malpractice lawsuit arising from a false negative interpretation. In particular, we show how combining double-read workflows, documentation practices, and the inclusion of AI error rates can reduce perceived liability. We also show that, when a double-read workflow is used, changing an initially correct interpretation to an incorrect one incurs greater liability for the radiologist than being incorrect in both the initial and final interpretations. These findings underscore the need to address cognitive biases that will undoubtedly arise at the human-AI interface.

*Implications of all the available evidence:* Optimizing the integration of AI tools into radiology requires strategic attention to workflow redesign, as combinations of features can collectively affect perceived liability, likely through well-known cognitive biases. Based on the results of our study, we propose one workflow that can mitigate a radiologist’s risk of legal liability. Moving forward, clinical practices and stakeholders should remain cognizant of these factors as they work toward building sustainable AI-physician systems.

## INTRODUCTION

Artificial intelligence (AI) is rapidly gaining momentum in medicine, particularly in diagnostic radiology.^1^ As a specialty, radiology is uniquely positioned to adopt AI for core tasks, as its primary clinical material is already in digital form. There is also an impetus for early adoption of AI tools in the field, driven by the ongoing demand-capacity mismatch, where the demand for imaging services due to rapid growth in imaging volumes overwhelms the capacity of interpretive services amid a worldwide shortage of radiologists. ^2^ AI-based solutions that aim to increase radiologists’ efficiency and reduce workloads span a range of applications from image triage to lesion detection.^3, 4^ In this way, AI in radiology has become a litmus test for the risks and benefits of delegating medical decision-making to AI models with varying levels of human-physician oversight.

Critically, this must be balanced with preserving diagnostic performance. Ideally—and perhaps, probably—the incorporation of AI will improve diagnostic performance, though it may have counterproductive effects, such as generating excessive false-positive detections that require further workup and additional imaging.^5^ This is especially relevant in settings with low disease prevalence, where even highly sensitive and specific AI systems will generate high rates of false-positive detections.^6, 7^ Moreover, as practices redesign radiology workflows to incorporate AI tools, they face several obstacles: technical interoperability barriers; human workforce integration; and, of central importance to this paper, questions about malpractice liability when patients are harmed by diagnostic errors made by a radiologist using AI. The spectre of legal liability has long been a motivating force in physician behavior^8,9^ and workflow redesign.^8,10,11^ Since AI error, like human error, is inevitable, whether radiologists using AI have met the legal standard of care will continue to be a paramount question. And while there are important non-legal costs to consider—personnel and overhead, for example—legal costs will attract stakeholders’ attention because they are sizeable and unpredictable.

Recent work has explored how these human-AI factors intersect in practice. For instance, one study showed that false-positive AI outputs can mislead radiologists into making errors that they would not have made without AI, likely due to a combination of anchoring and/or automation biases.^12^ Moreover, consistent with the adaptive bias from error management theory^13^ and loss aversion from prospect theory,^14,15^ this phenomenon was exacerbated when radiologists believed AI results would be documented in the patient record. Radiologists have also expressed concerns about legal liability repercussions when they disagreed with the AI output.^6^ Thus, besides AI standalone performance, workflow integration and documentation practices also shape concerns related to malpractice risk.

Radiologists’ fears that AI will increase their liability risk are not unfounded. In a jury simulation experiment, hypothetical jurors were asked to indicate if they would side with a plaintiff or defendant (radiologist) after the defendant failed to detect an abnormality that ultimately led to injury or death. When the AI output correctly detected the abnormality, jurors were more likely to side with the plaintiff than when the AI output also failed to detect the abnormality or in the case where no AI system was used.^16^ Thus, the study revealed an “AI penalty” in the eyes of the jury. That is, AI was found to increase a radiologist’s perceived liability when AI detected an abnormality that a radiologist missed, but provided no corresponding protection from liability when AI also failed to detect the abnormality.

Empirical work has identified factors that can mitigate this liability. Of note, providing mock jurors with information about the AI false discovery rate (FDR) and false omission rate (FOR) significantly decreased the likelihood that jurors ruled against the radiologist.^16^ This finding was consistent with results of another prior study, which suggested that women who were asked to imagine being ultimately diagnosed with breast cancer, and whose mammogram was originally flagged as abnormal by AI but interpreted as showing no abnormality by the radiologist, were less likely to consider a medical malpractice lawsuit if made aware of AI’s FDR.^17^ Further experimental work demonstrated that workflow design also influences perception of liability. When a radiologist reviewed a case using a single-read workflow—interpreting the image after viewing AI output—perceived liability for a false negative interpretation was substantially greater compared to when a double-read workflow was used, where the radiologist provided an independent tentative interpretation before consulting AI, then a final interpretation after viewing AI feedback.^18^ In the double-read condition, the radiologist failed to detect the abnormality during both the tentative and final interpretation.

The present study builds upon this prior research by examining how multiple dimensions of AI implementation, including workflows and contextual features, collectively affect perceptions of legal liability. In doing so, we aim to improve stakeholders’ ability to anticipate medico-legal outcomes when radiologists use AI tools for decision-making in cases where errors result in patient harm. Some of the central questions that we seek to answer in this study are listed in Box 1. Specifically, we compare the combined effects of single-read versus double-read workflows, radiologists’ awareness of AI error rates, and documentation of radiologists’ interpretations before and after AI consultation on perceived legal liability. We also compared conditions to a “no AI” control. This study aims to identify modifiable aspects of AI implementation that may mitigate unintended malpractice risk to radiologists. Our hypotheses are listed in Table 1.

**Table 1.**
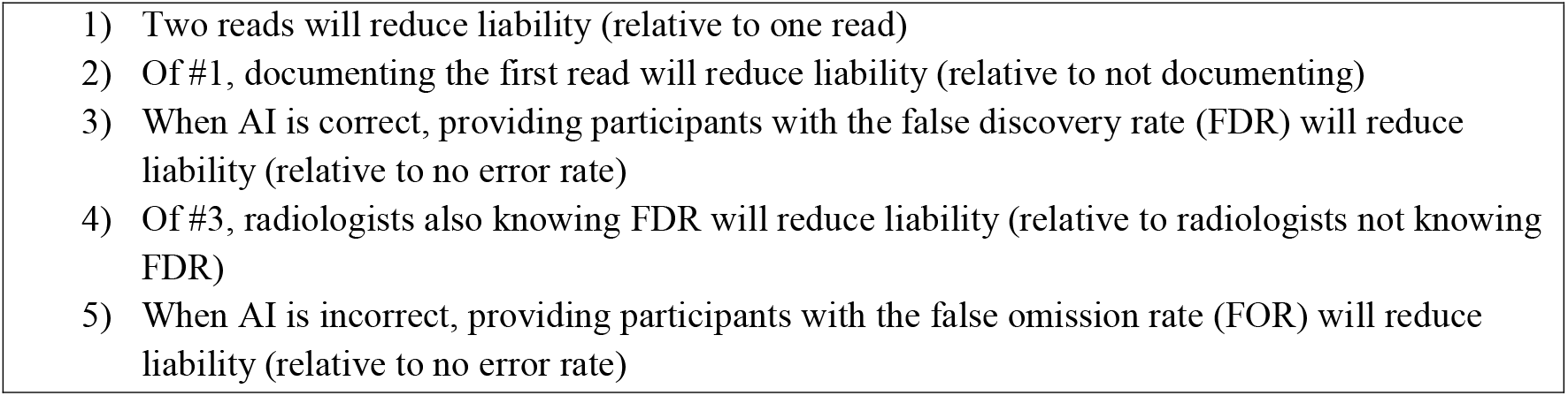
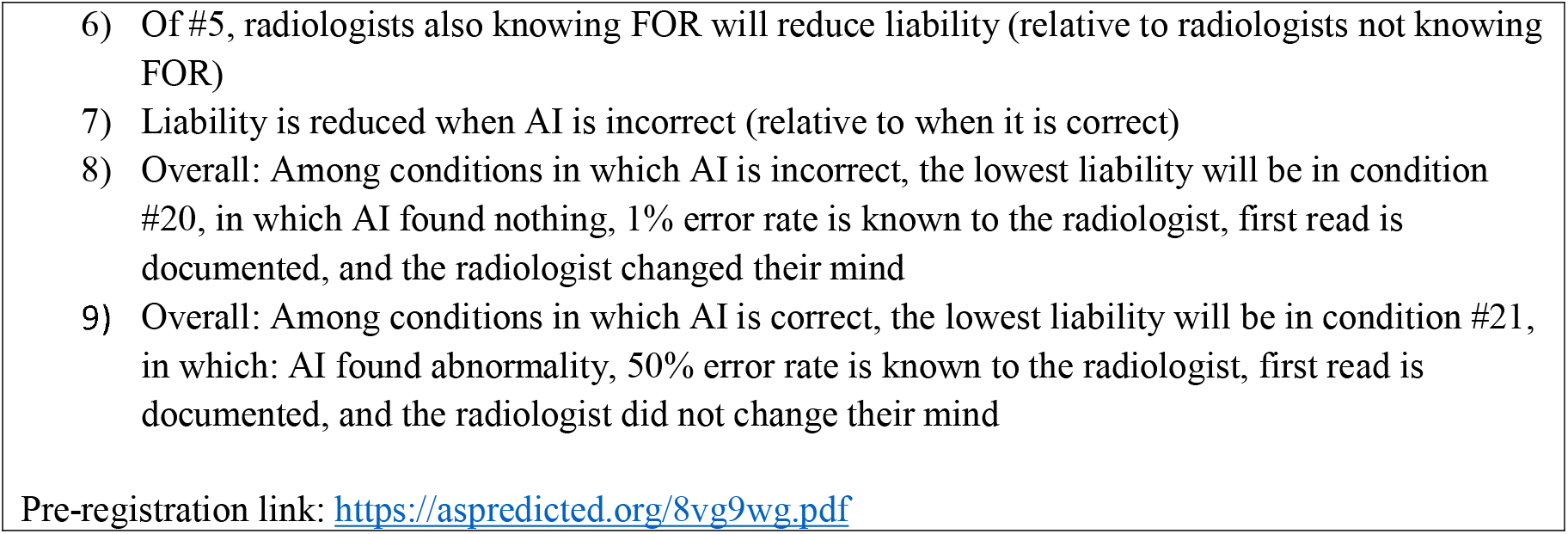
Hypotheses.

### Box 1.

Questions regarding legal liability and AI in radiology

1. Will jurors be less favorable to doctors for disagreeing with AI when AI is correct?
2. Will jurors be more favorable to doctors for agreeing with AI when AI was incorrect?
3. Will jurors prefer that a doctor knows the AI determination before reading an image versus after?
  a. If after, will juror perceptions differ according to whether or not the radiologist changed their mind between the two interpretations?

## METHODS

### Participants

Participants were recruited via the online platform Prolific (London, UK). To emulate a real-world jury, our inclusion criteria were broad: participants were eligible if they were 18 to 89 years old, resided in the United States, and spoke English as their primary language. A total of n=5,555 participants were recruited; n=545 responses were dropped because of non-completion, ReCAPTCHA score failures (<0.80), and/or potential duplicate responses (flagged by Qualtrics). Using a conservative approach, of the remaining 5,010 responses, 2,663 were removed because they missed one or more comprehension checks or manipulation checks, leaving a total of n=2,347 responses. Of the 2,347 respondents, 1,331 (56.7%) identified as female, 1,818 identified as White (77.5%), 249 (10.6%) as Black, and 150 (6.4%) as Asian, with most respondents having finished high school and the majority having at least some college or more (see Supplemental Table 1).

### Procedure

The Brown University Health Institutional Review Board waived ethical approval for this work (IRB # 2323232). The clinical vignette used for this experiment parallels that of prior studies.^16,18^ Online volunteers who opted into the study and met the eligibility criteria were provided with an informed consent page. If they consented, they were directed to a Qualtrics (Provo, UT) survey where they were randomized to one of twenty-two conditions. Participants were compensated $1.61 for completing the survey and were prevented from participating more than once.

### Vignette Design and Experimental Conditions

Eligible participants were presented with a hypothetical case in which a radiologist had failed to identify a brain bleed on a CT scan of a stroke patient, leading to the administration of tissue plasminogen activator (tPA), a blood-thinning medication. Participants were told this treatment exacerbated bleeding into the brain, leading the patient to suffer irreversible brain damage. Participants, much like actual jurors, were tasked with deciding whether the radiologist had met their duty of care to the patient. The vignettes are presented in Supplementary Materials.

Each participant was randomized to one of twenty-two conditions representing varying radiologist workflows and levels of AI use (Box 2). The control—condition #1—had no mention of AI. The remaining twenty-one conditions followed a 7 x 3 design consisting of seven distinct radiologist-AI workflow scenarios. The seven workflows differed in whether:

1. the radiologist interpreted the image once—after seeing AI output, or twice—once before and once after seeing the AI output (i.e., Reviewed Once vs. Reviewed Twice),
2. the radiologist’s initial interpretation was documented or undocumented (i.e. Doc vs. UnDoc; Reviewed Twice conditions only)
3. the AI output detected the abnormality or failed to detect the abnormality (i.e. AI Detected vs. AI Failed to Detect), and
4. the radiologist changed their initial interpretation after viewing AI output or not (i.e. changed vs. not changed; Reviewed Twice conditions only).

As summarized in Box 2 and Table 2, two workflows—conditions #2 and 3—involved a single-read (Reviewed Once) in which the radiologist reviewed the AI output and either disagreed or agreed with its interpretation as positive for a brain bleed. Five workflows—conditions #4-8—involved a double-read (Reviewed Twice) in which the radiologist made an initial interpretation, and then viewed the AI output to make a final determination. Among these five conditions, two workflows—conditions #4 and 5— involved a second read following an undocumented initial radiologist interpretation (UnDoc), with AI either detecting an abnormality or failing to detect an abnormality, respectively; and three workflows— conditions #6-8—involved a second read following a documented initial interpretation (Doc), including one in which the radiologist reversed (henceforth “Changed”) an initially correct interpretation after viewing AI output where AI failed to detect the abnormality (condition 6) and two in which the radiologist maintained an initial incorrect interpretation where AI either detected (i.e. condition 7) or failed to detect (i.e. condition 8) the abnormality.

**Table 2.**
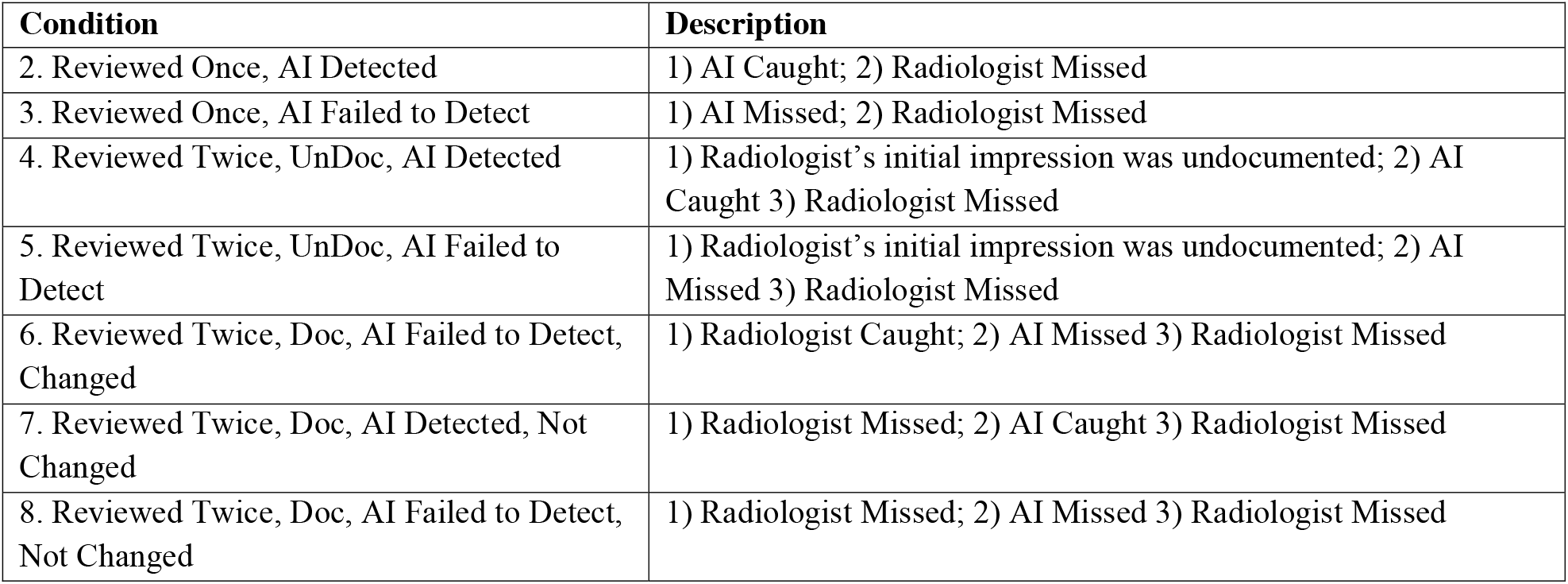
Description of Basis conditions #2-8.

| Condition | Description |
| --- | --- |
| 2. Reviewed Once, AI Detected | 1) AI Caught; 2) Radiologist Missed |
| 3. Reviewed Once, AI Failed to Detect | 1) AI Missed; 2) Radiologist Missed |
| 4. Reviewed Twice, UnDoc, AI Detected | 1) Radiologist's initial impression was undocumented; 2) AI Caught 3) Radiologist Missed |
| 5. Reviewed Twice, UnDoc, AI Failed to Detect | 1) Radiologist's initial impression was undocumented; 2) AI Missed 3) Radiologist Missed |
| 6. Reviewed Twice, Doc, AI Failed to Detect, Changed | 1) Radiologist Caught; 2) AI Missed 3) Radiologist Missed |
| 7. Reviewed Twice, Doc, AI Detected, Not Changed | 1) Radiologist Missed; 2) AI Caught 3) Radiologist Missed |
| 8. Reviewed Twice, Doc, AI Failed to Detect, Not Changed | 1) Radiologist Missed; 2) AI Missed 3) Radiologist Missed |

Each workflow was presented in three ways: no AI error information, AI error information provided to participants but participants were explicitly told it was *unknown* to the radiologist (i.e., Unaware), and AI error information provided to jurors while explicitly told it was *known* to the radiologist (i.e., Aware). When error information was provided, the type of information was dependent on the AI output: a false discovery rate (FDR) was reported when AI flagged a bleed (AI Detected conditions), and a false omission rate (FOR) was reported when AI did not flag a bleed (AI Failed to Detect conditions). FDR and FOR values were rounded into whole numbers for interpretability assuming a sensitivity and specificity of 90% and a prevalence of 10%, consistent with a previous study.^16^

### Measures

#### Culpability

As with the prior studies, radiologist (defendant) culpability was assessed with a single yes/no question: “Did the radiologist meet their duty of care to the patient?” A response of “no” indicates siding with the plaintiff (i.e., finding the radiologist liable); a response of “yes” indicates siding with the defendant (i.e., not finding the radiologist liable). Results are interpreted as the percentage of participants who sided with the plaintiff. Prior to answering this item, participants were told, “Radiologists, as doctors, owe a duty of care to the patients for whom they are reviewing imaging. This means that, when they review images, radiologists must use the level of care, skill, and knowledge ordinarily used by radiologists under the circumstances of the case you just read about.”

### Comprehension and Manipulation Checks

Three comprehension check items were included to verify participants’ understanding of the vignette content and five manipulation checks (depending on the experimental condition) were included to verify participants understood the manipulations (see Supplementary Materials).

#### Box 2.

Experimental conditions

1. No AI (control) **\*Basis (2-8)**
2. Reviewed Once, AI Detected
3. Reviewed Once, AI Failed to Detect
4. Reviewed Twice, UnDoc, AI Detected
5. Reviewed Twice, UnDoc, AI Failed to Detect
6. Reviewed Twice, Doc, AI Failed to Detect, Changed
7. Reviewed Twice, Doc, AI Detected, Not changed
8. Reviewed Twice, Doc, AI Failed to Detect, Not changed **\*Same as 2-8, but Radiologist is Unaware of Error Rates (i.e. Unaware Conditions)**
9. Reviewed Once, AI Detected, FDR-Unaware
10. Reviewed Once, AI Failed to Detect, FOR-Unaware
11. Reviewed Twice, UnDoc, AI Detected, FDR-Unaware
12. Reviewed Twice, UnDoc, AI Failed to Detect, FOR-Unaware
13. Reviewed Twice, Doc, AI Failed to Detect, Changed, FOR-Unaware
14. Reviewed Twice, Doc, AI Detected, Not changed, FDR-Unaware
15. Reviewed Twice, Doc, AI Failed to Detect, Not changed, FOR-Unaware **\*Same as 9-15 but Radiologist is Aware of Error Rates (i.e. Aware Conditions)**
16. Reviewed Once, AI Detected, FDR-Aware
17. Reviewed Once, AI Failed to Detect, FOR-Aware
18. Reviewed Twice, UnDoc, AI Detected, FDR-Aware
19. Reviewed Twice, UnDoc, AI Failed to Detect, FOR-Aware
20. Reviewed Twice, Doc, AI Failed to Detect, Changed, FOR-Aware
21. Reviewed Twice, Doc, AI Detected, Not changed, FDR-Aware
22. Reviewed Twice, Doc, AI Failed to Detect, Not changed, FOR-Aware

### Power analysis

Power to detect was set at 90%, assuming a 5% alpha value using a Pearson Chi-square Test for Proportion Difference with effect size and variance estimates from two previous studies demonstrating the effect of reducing liability; specifically, when including FDR with AI detection compared to not including the FDR (0.49 vs. 0.73)^16^ and providing AI results only after an initial read without AI vs. simultaneously (0.53 vs. 0.75),^18^ yielding a minimum sample size of n=69 and n=80, respectively. In order to anticipate loss of sample due to comprehension and manipulation check failures, and to power for hypothesis with no previous estimates and for false discovery correction, we aimed at collecting n=130 per condition, or n=2,860.

### Statistical analysis

All modeling was conducted using SAS Software 9.4 (Cary, NC). Duty of care being met or not (1/0) was examined using generalized linear modeling assuming a binary distribution with the GLIMMIX procedure. Specific hypotheses were examined using planned comparisons. Alpha was established a priori at 0.05, and all interval estimates were calculated for 95% confidence. Exploratory comparisons were adjusted using FDR correction at q=0.05.

## RESULTS

### Hypothesized Results

As illustrated in Figure 1 and Tables 3 and 4, perception of duty of care varied greatly between conditions, p<0.0001. As seen in Figure 1 and Tables 3 and 4, as hypothesized, liability was reduced when:

**Figure 1.**
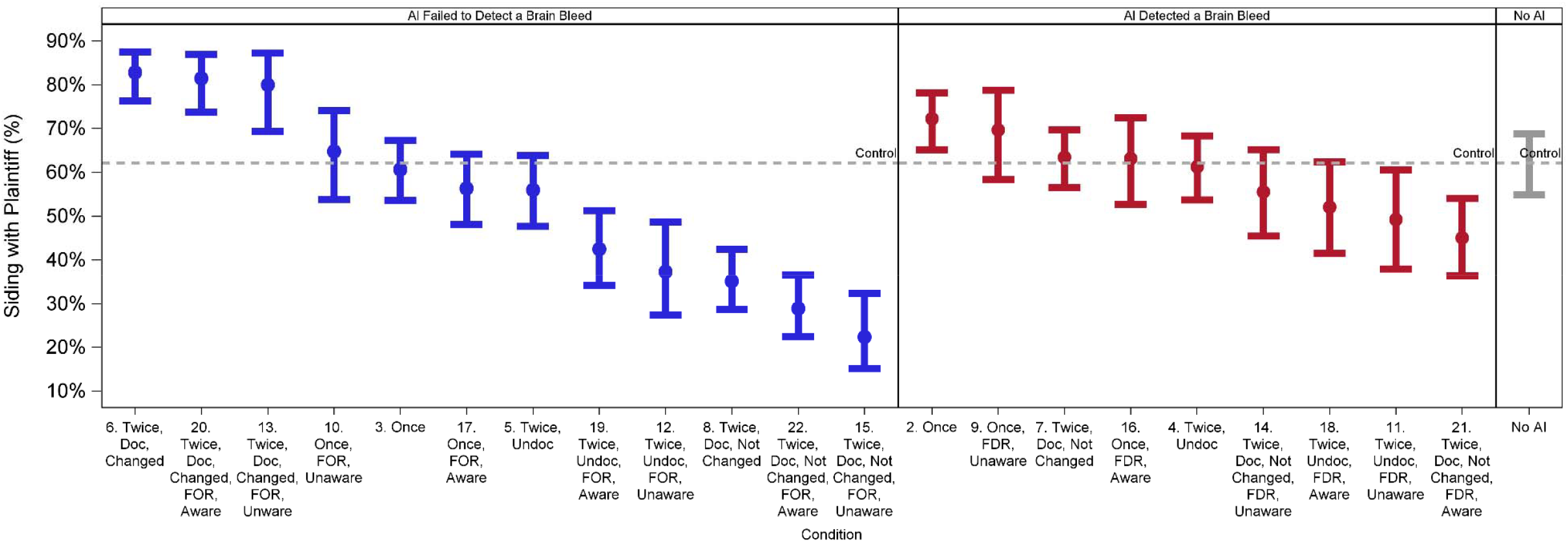
Mean with 95% CIs by experimental condition. X-axis denotes the experimental condition: Blue corresponds to when AI failed to detect a brain bleed, Red when AI detected a brain bleed, and Gray is the control condition, No AI. Y-axis is percentage agreeing with plaintiff. “Once” refers to the radiologist interpreting imaging coterminously with AI and “Twice” refers to the radiologist interpreting imaging first without AI then with AI. “FOR” refers to the False Omission Rate and “FDR” refers to the False Discovery Rate. “Aware” refers to the radiologist being aware of the error rate and “Unaware” refers to the radiologist being unaware of the error rate. “Doc” refers to the initial interpretation without AI being documented and “UnDoc” refers to the initial interpretation being undocumented. “Changed” refers to the initial impression being changed (i.e., positive to a negative) and “Unchanged” refers to the initial impression being unchanged (i.e., a negative stayed a negative). No AI refers to the control condition where no AI was used. For all conditions, the final radiologist interpretation was no evidence of brain bleed.

**Table 3.** Mean and 95% CI by Experimental Condition.

| AI Result |  | Condition | Siding with plaintiff (%) | LL (%) | UL (%) |
| --- | --- | --- | --- | --- | --- |
| **Failed to Detect** | **Changed** | 6. Reviewed Twice, Doc, Changed | 82.8 | 75.5 | 88.3 |
|  |  | #20. Reviewed Twice, Doc, Changed, FOR-Aware | 81.5 | 73.0 | 87.7 |
|  |  | 13. Reviewed Twice, Doc, Changed, FOR-Unaware | 80.0 | 68.5 | 88.0 |
|  |  | 10. Reviewed Once, FOR-Unaware | 64.8 | 53.1 | 75.0 |
|  |  | 3. Reviewed Once, AI Failed to Detect | 60.7 | 52.8 | 68.0 |
|  |  | 17. Reviewed Once, FOR-Aware | 56.3 | 47.3 | 64.9 |
|  |  | 5. Reviewed Twice, UnDoc, AI Failed to Detect | 55.9 | 46.9 | 64.6 |
|  |  | 19. Reviewed Twice, UnDoc, FOR-Aware | 42.5 | 33.4 | 52.0 |
|  |  | 12. Reviewed Twice, UnDoc, FOR-Unaware | 37.3 | 26.6 | 49.4 |
|  | **Not changed** | 8. Reviewed Twice, Doc, Not changed | 35.1 | 27.9 | 43.2 |
|  |  | 22. Reviewed Twice, Doc, Not changed, FOR-Aware | 28.9 | 21.7 | 37.3 |
|  |  | 15. Reviewed Twice, Doc, Not changed, FOR-Unaware | 22.4 | 14.4 | 33.1 |
| **Detected** |  | 2. Reviewed Once, | 72.2 | 64.4 | 78.9 |
|  |  | 9. Reviewed Once, FDR-Unaware | 69.7 | 57.6 | 79.6 |
|  | **Not changed** | 7. Reviewed Twice, Doc, Not changed | 63.4 | 55.8 | 70.4 |
|  |  | 16. Reviewed Once, FDR-Aware | 63.2 | 51.8 | 73.2 |
|  |  | 4. Reviewed Twice, UnDoc, | 61.3 | 52.9 | 69.1 |
|  | **Not changed** | 14. Reviewed Twice, Doc, Not changed, FDR-Unaware | 55.6 | 44.6 | 66.0 |
|  |  | 18. Reviewed Twice, UnDoc, FDR-Aware | 52.1 | 40.7 | 63.2 |
|  |  | 11. Reviewed Twice, UnDoc, FDR-Unaware | 49.2 | 37.2 | 61.4 |
|  | **Not changed** | **21. Reviewed Twice, Doc, Not changed, FDR-Aware** | **45.0** | **35.6** | **54.8** |
| **No AI** |  | 1. No AI (control) | 62.2 | 54.1 | 69.6 |

Note: Mean and 95% CIs displayed for all conditions. Outcome is siding with the plaintiff (finding radiologist liable). LL and UL are Lower Limit and Upper Limit, respectively. Condition Number reflects Box 2, and the order of conditions reflects Figure 1. FDR=false discovery rate; FOR = false omission rate
\*Confirmed hypothesis #9
#Failed to confirm hypothesis #8

**Table 4.**
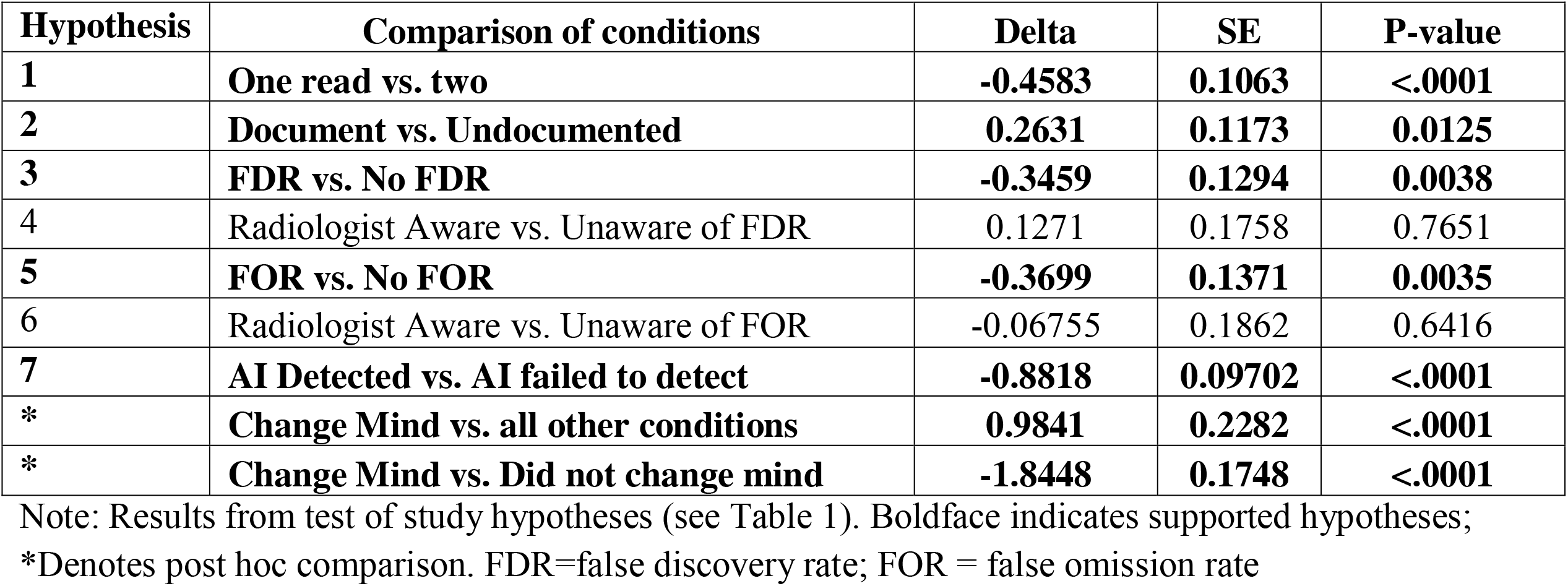
Planned contrasts for hypotheses.

| **Hypothesis** | **Comparison of conditions** | **Delta** | **SE** | **P-value** |
| --- | --- | --- | --- | --- |
| **1** | **One read vs. two** | **-0.4583** | **0.1063** | **<.0001** |
| **2** | **Document vs. Undocumented** | **0.2631** | **0.1173** | **0.0125** |
| **3** | **FDR vs. No FDR** | **-0.3459** | **0.1294** | **0.0038** |
| 4 | Radiologist Aware vs. Unaware of FDR | 0.1271 | 0.1758 | 0.7651 |
| **5** | **FOR vs. No FOR** | **-0.3699** | **0.1371** | **0.0035** |
| 6 | Radiologist Aware vs. Unaware of FOR | -0.06755 | 0.1862 | 0.6416 |
| **7** | **AI Detected vs. AI failed to detect** | **-0.8818** | **0.09702** | **<.0001** |
| **\*** | **Change Mind vs. all other conditions** | **0.9841** | **0.2282** | **<.0001** |
| **\*** | **Change Mind vs. Did not change mind** | **-1.8448** | **0.1748** | **<.0001** |

Note: Results from test of study hypotheses (see Table 1). Boldface indicates supported hypotheses;
\*Denotes post hoc comparison. FDR=false discovery rate; FOR = false omission rate

*Hypothesis 1*. The radiologist read the case initially without AI and then subsequently with AI compared to the radiologist reading the case only once with AI, p<0.0001.

*Hypothesis 2*. The radiologist’s initial impression without AI was documented relative to it not being documented, p=0.0125.

*Hypothesis 3*. The AI result was provided with the FDR when AI correctly flagged the case compared to without the FDR, p=0.0038.

*Hypothesis 5*. The AI result was provided with the FOR when AI incorrectly failed to flag the case compared to without the FOR, p= 0.0035.

*Hypothesis 7*. When AI was also incorrect (failed to detect) compared to when AI was correct (detected), p<0.0001.

In addition, per hypothesis 9, when AI detected an abnormality, the combination of variables that achieved the lowest perceived legal liability for the physician was when [1] the AI’s FDR was known to the radiologist, [2] the radiologist’s impression of the initial read without AI was documented, and [3] the radiologist did not change their mind after discovering the AI had flagged the case (M=45%).

We failed to find evidence supporting hypotheses 4, 6, or 8. Specifically, there was no evidence that the radiologist being Aware (versus not being Aware) of the FDR (p=0.7651) or the FOR (p=0.6416) reduced liability (hypotheses 4 and 6, respectively). In addition, our results did not support the hypothesized pattern for what condition achieved the lowest liability when AI failed to detect the pathology. That combination (FOR was known to the radiologist, the radiologist’s impression of the initial read without AI was documented, and the radiologist changed their mind), was actually among the highest levels of liability (M=81.5%).

### Exploratory Results

As illustrated in Figure 1 and Tables 3 and 5, the lowest liability was achieved in condition #15, where AI failed to detect an abnormality, a double-read workflow was used, the radiologist’s impression of the initial read without AI was documented, and the radiologist was unaware of the FOR (M=22.4%). The second lowest liability was seen with this same combination, except the radiologist was Aware of the FOR (condition #22, M=28.9%).

**Table 5.** Exploratory Comparisons with No AI condition.

| Condition | Control | Delta | SE | P-value | FDR | +/- |
| --- | --- | --- | --- | --- | --- | --- |
| **AI Failed to Detect** |  | **log-odds scale** |  |  | **q=0.05** |  |
| 6. Reviewed Twice, Doc, Changed | No AI | 1.0776 | 0.285 | 0.0002 | **0.00105** | + |
| 20. Reviewed Twice, Doc, Changed, FOR-Aware | No AI | 0.9852 | 0.3002 | 0.001 | **0.00350** | + |
| 13. Reviewed Twice, Doc, Changed, FOR-Unaware | No AI | 0.8899 | 0.3534 | 0.0119 | **0.02777** | + |
| 10. Reviewed Once, FOR-Unaware | No AI | 0.1133 | 0.3008 | 0.7064 | 0.87261 | + |
| 3. Reviewed Once, AI Failed to Detect | No AI | -0.06402 | 0.2361 | 0.7863 | 0.88410 | - |
| 17. Reviewed Once, FOR-Aware | No AI | -0.243 | 0.2508 | 0.3327 | 0.43667 | - |
| 5. Reviewed Twice, UnDoc, AI Failed to Detect | No AI | -0.258 | 0.2512 | 0.3045 | 0.43667 | - |
| 19. Reviewed Twice, UnDoc, FOR-Aware | No AI | -0.8006 | 0.2595 | 0.0021 | **0.00630** | - |
| 12. Reviewed Twice, UnDoc, FOR-Unaware | No AI | -1.0152 | 0.3042 | 0.0009 | **0.00350** | - |
| 8. Reviewed Twice, Doc, Not changed | No AI | -1.1095 | 0.2416 | <.0001 | **0.00070** | - |
| 22. Reviewed Twice, Doc, Not changed, FOR-Aware | No AI | -1.3964 | 0.2583 | <.0001 | **0.00070** | - |
| 15. Reviewed Twice, Doc, Not changed, FOR-Unaware | No AI | -1.7408 | 0.3233 | <.0001 | **0.00070** | - |
| **AI Detected** |  |  |  |  |  |  |
| 2. Reviewed Once, AI Detected | No AI | 0.4591 | 0.2517 | 0.0683 | 0.14343 | + |
| 9. Reviewed Once, FDR-Unaware | No AI | 0.3365 | 0.317 | 0.2886 | 0.43667 | + |
| 7. Reviewed Twice, Doc, Not changed | No AI | 0.05361 | 0.2345 | 0.8192 | 0.88410 | + |
| 16. Reviewed Once, FDR-Aware | No AI | 0.04256 | 0.292 | 0.8841 | 0.88410 | + |
| 4. Reviewed Twice, UnDoc, AI Detected | No AI | -0.03591 | 0.2439 | 0.883 | 0.88410 | - |
| 14. Reviewed Twice, Doc, Not changed, FDR-Unaware | No AI | -0.2733 | 0.2806 | 0.3301 | 0.43667 | - |
| 18. Reviewed Twice, UnDoc, FDR-Aware | No AI | -0.4142 | 0.2892 | 0.1522 | 0.26635 | - |
| 11. Reviewed Twice, UnDoc, FDR-Unaware | No AI | -0.5282 | 0.3037 | 0.0821 | 0.15674 | - |
| 21. Reviewed Twice, Doc, Not changed, FDR-Aware | No AI | -0.6971 | 0.2629 | 0.0081 | **0.02126** | - |

+/-: higher (+) or lower (-) than control. FDR=false discovery rate; FOR = false omission rate.
Condition Number reflects Box 2, and the order of conditions reflects Figure 1.

The highest liability was observed in conditions #6, 13, and 20, where the radiologist had correctly documented an initial abnormal read without AI, learned that AI did not flag the case, and changed their mind to decide the case was normal. These high rates of liability occurred without mention of FOR (condition #6, 82.8%), when participants were provided the FOR knowing the radiologist was unaware of it (condition #13, 80.0%), and when participants were given the FOR and told the radiologist also knew the FOR (condition #20, 81.5%). Liability was higher in all three of these conditions compared to the scenario in which the radiologist did not change their mind (p <0.0001), as well as all other AI conditions (p <0.0001) and the control condition (p=0.0002, p=0.0119, and p=0.001, respectively).

Finally, conditions #8, 12, 15, 19, 21, and 22 all achieved lower liability than the “no AI condition,” while all other conditions failed to differ from the no AI control.

## DISCUSSION

In our study, several factors were identified that modified a radiologist’s perceived liability for a false-negative interpretation error when AI is used. Radiologist liability was reduced when AI also provided a false negative interpretation versus a true positive interpretation, when jurors were told the AI error rates, and when radiologists interpreted a case twice versus once. Thus, the present study replicates these prior findings^16,18^ and extends this line of research by also examining whether the following factors modified perceived liability: 1) A radiologist’s awareness of AI’s FDR and FOR, 2) whether the radiologist documented their initial interpretation prior to reviewing AI output in a double-read workflow, and 3) whether a radiologist changed their mind in a double-read workflow.

Our results support the conclusion that reducing legal liability is best achieved when the radiologist reads the case twice and is aware of the AI error rates. In these double-read conditions, the radiologist first documented an initial impression without knowing the AI output, then again with the AI output accompanied by its FDR/FOR. Critically, however, our results indicate that if a radiologist documents an abnormality on their initial read and then changes their mind after seeing that AI failed to alert the case, they will incur the maximum perception of liability from a jury—an “erroneous change penalty.” In short, when radiologists fail to detect an abnormality, they are less likely to be found liable when they *consistently* fail to detect it, as radiologists had higher perceived liability when they changed their mind (i.e., initially detecting the abnormality) versus when they did not change their mind (i.e., not initially detecting the abnormality).

### Psychological Theory for the Erroneous-Change Penalty

Research from the domains of psychological and cognitive sciences provides some possible explanations for why the erroneous change penalty occurred. Just as cognitive biases can shape the behavior of radiologists themselves,^10^ our work suggests that it can also modify the perception of jurors who are evaluating radiologists’ errors. Kruger and colleagues^19^ suggest that there is a “first instinct penalty.” People have an inflated belief in the accuracy of their first instinct, and are harsher on both themselves and others when a correct response is changed to an incorrect response, versus when an incorrect response is not preceded by the correct answer.^19^ Similarly, an incorrect answer that is a “near miss” is viewed more negatively than an incorrect answer that is “far miss.” ^20,21^

Another cognitive bias that could explain the erroneous change penalty is the “omission bias,” which is the tendency to judge harmful outcomes resulting from inaction as less blameworthy than equivalent harms resulting from action.^22^ One meta-analysis of 21 studies where the same harm occurred due to experimentally manipulated errors of omission or commission found that errors of commission were judged as more immoral and blameworthy (*g*=0.71 and 0.32, respectively).^23^ This is consistent with “regret aversion,” which suggests that people do not rationally optimize for the decision that yields the maximum expected value, but instead are motivated to avoid making decisions that could cause future regret when facing uncertainty.^24^ For instance, Engelstein^25^ describes an experiment in which volunteers must pick one of two cups; one cup, at random, corresponds with a monetary prize while the other corresponds with no prize. Participants were also given the option to change their selection before finding out whether they picked the cup that yields a monetary reward. Even though they have an equal chance of winning (50%) regardless of whether or not they choose to switch, only about 10% switch, presumably because switching and being wrong would induce regret. Importantly, even when participants are told that their reward will triple if they correctly switch, only about half of participants switched despite the fact that switching is the rational decision from a profit-maximizing standpoint.

These studies relate to the current experiment in several key ways. First, because the radiologist in the “change” conditions shifts from the correct answer to the incorrect answer, it may evoke in jurors Kruger’s first instinct penalty.^19^ Along similar lines, jurors who saw the radiologist shift from correct to incorrect answer might have been more likely than jurors who saw only an incorrect answer to characterize the radiologist’s mistake as a near miss, which could make those jurors to be more likely to engage in critical counterfactual thinking. As such, the cognitive biases outlined by Kruger, et al., and by Roese may lead mock jurors to judge a radiologist who changes their mind from a correct to an incorrect interpretation as committing a more egregious error than a radiologist who is always incorrect.^19,20^ Moreover, when a radiologist initially interprets a study correctly but then revises their interpretation, making it incorrect, jurors may perceive this as an error of commission: the radiologist actively changed their judgment, and in doing so, caused harm. Conversely, when a radiologist does not revise an initially incorrect interpretation, the error may be perceived as a simple failure to detect an abnormality—a passive lapse that is instead an error of omission.

Researchers have suggested that these aforementioned cognitive biases may occur due to the cognitive ease with which one can access counterfactuals. In reviewing related work, Kahneman and Miller^26^ describe this as follows: “[A] negative fate for which a more positive contrast is highly available is [seen as] worse or more unfair than one for which there is no highly available positive alternative.”

Near misses may be perceived as worse than far misses because it is easier to imagine the alternative of “correct” or “no harm” in the former scenario. In an early study highly relevant to our experiment, mock jurors awarded more compensation to the victim of a plane crash that occurred ¼ mile from the final destination than to one 75 miles from it.^26^ Just as it is easier to imagine a safe return home when the accident occurs at the very end of the flight, it is also easier to imagine the alternative of “no error” when evaluating an error of omission versus commission.^27^ This is supported by the empirical finding that errors of commission are viewed as more causally linked to the same harm than errors of omission.^28^ As such, in our experiment, the counterfactual of a true positive (versus a false negative) radiologist interpretation may be more readily accessible cognitively when the error is one of commission (no change) versus omission (change), thus making the harm more salient in the latter scenario.

Regardless of the exact mechanism, our study underscores the fact that the radiologist and AI interpretations and their sequence were highly relevant evidence for the perception of legal liability. The documentation of the initial interpretation in a double-read scenario largely reduced perceptions of liability, while changing one’s mind increased liability. Thus, the very documentation that protects a radiologist can also expose them to liability depending on whether the initial and final interpretation are the same or different.

## Limitations

This study is not without limitations. As with previous studies, the vignettes presented to participants were likely less detailed than what would be presented to a real jury, limiting ecological validity. Furthermore, participants responded independently and were unable to deliberate together like real jurors. However, both of these factors suggest that the magnitude of the effects observed in this study might be even greater in a real-world context, as real jurors would likely process information more carefully and have the opportunity to exchange ideas.

While our sampling strategy attempted to capture a broad range of the US adult population, Prolific participants may not be representative of the US population as a whole, and we did not screen participants for jury eligibility criteria such as citizenship status and criminal history; nonetheless, these sampling biases, if they exist, would apply across all conditions because assignment was random, and thus do not jeopardize the interpretation of between-condition differences.

A final limitation is that neither the defendant nor the plaintiff questioned the authenticity of the documentation, which was necessary from an experimental standpoint. Although beyond the scope of this study, in real-world settings, if documentation can reduce or enhance perceptions of liability, then one party may be incentivized to dispute or question its accuracy, and the documentation itself could be placed into question.^29^

### Proposed Workflow

With these results in mind, we propose a workflow to mitigate legal risk, as illustrated in Figure 2: (1) the radiologist reads all cases initially without knowing AI results, and their initial (human-only) interpretation is fully documented. (2a) On this initial (human-only) review, if the radiologist decides that there is evidence of an abnormality, then they would not review the AI output—as they are disincentivized from changing their mind between the initial and final interpretation. (2b) Only in cases when the radiologist believes there to be no evidence of an abnormality on the initial read should the AI output, along with the FDR/FOR, be reviewed. As a secondary advantage, by only double-reading cases identified as normal during the initial interpretation, the total radiologist caseload is reduced, which is important given the ongoing radiologist shortage.^30^

**Figure 2.**
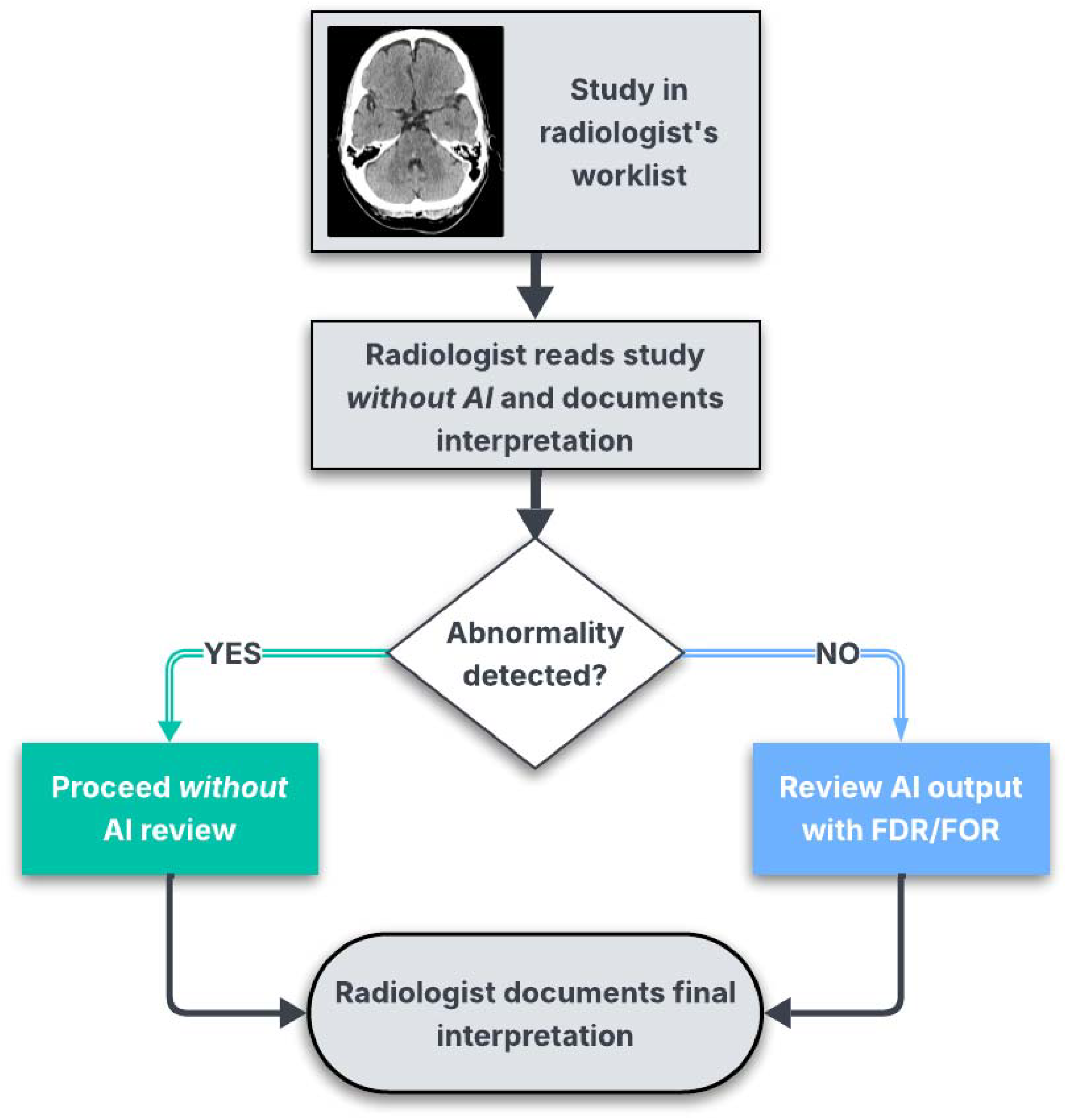
Optimal workflow for mitigating legal liability.

## Data Availability

All data produced in the present study are available upon reasonable request to the authors

## Supplemental Table

**Supplemental Table 1.** Demographics.

| **Sex** | **Count** | **Percentage** |
| --- | --- | --- |
| Female | 1331 | 56.8 |
| Male | 1012 | 43.2 |
| **Race** |  |  |
| Asian American | 150 | 6.4 |
| Black/African American | 249 | 10.6 |
| More than one race | 106 | 4.5 |
| Native Hawaiian/other | 5 | 0.2 |
| Pacific Islander<br>Indigenous/Native<br>American | 17 | 0.7 |
| White/Caucasian | 1818 | 77.5 |
| **Highest Education** |  |  |
| Associate | 232 | 9.9 |
| Bachelor's | 893 | 38.3 |
| Did not finish high school | 11 | 0.5 |
| Doctoral or professional<br>degree | 114 | 4.9 |
| High school diploma or<br>GED | 257 | 11.0 |
| Master's | 374 | 16.0 |
| Some college | 453 | 19.4 |
| Median (q1-q3) Income | \$74,000.00<br>(\$41,284.50-\$119,000.00) | |

